# Partnering for Progress: Lessons Learned from a Mental Health Assessment for Youth Living with HIV in India through Community-Based Participatory Research

**DOI:** 10.1101/2024.04.08.24304947

**Authors:** Siddhaparna Sannigrahi, Babu Seenappa, Prashant Laxmikanth, Suhas Reddy, Kacie Filian, Michael Babu Raj, Lakshmi Ganapathi, Anita Shet

## Abstract

YLHIV face diverse mental health challenges necessitating interventions informed by their lived experiences. Failure to do so can perpetuate a self-reinforcing cycle of misaligned and ineffective support, further exacerbating existing vulnerabilities. This study directly addresses this gap by using a community-based participatory research (CBPR) approach to empower Youth Living with HIV (YLHIV) in India to address their mental health challenges. YLHIV actively participated in adapting and implementing a culturally responsive mental health screening program. The study, incorporating CBPR principles at every step aimed to identify practical methods for integrating YLHIV voices in research, and showcase the value of YLHIV participation in co-creating and implementing impactful interventions. Six youth investigators aged 18-24, born with HIV and residing in southern Indian states underwent certification in human subjects’ protection and training in mental health screening and survey administration techniques. They actively shaped the research process by culturally adapting standardized mental health screening tools (PHQ-9, GAD-7) through iterative discussions with experts, drawing on their own perspectives. Following field assessments of the mental health screening tools among their peers, they documented their reflections in surveys and written essays. Youth investigators’ involvement improved the research process by optimizing tools, combating stigma, and facilitating reliable data collection. Beyond data collection, the youth investigators’ participation significantly boosted their own knowledge, self-confidence, and research skills. This study serves as an illustrative model of CBPR in mental health research among YLHIV, highlighting the importance of interactive training, continuous feedback mechanisms, and respectful youth engagement in fostering impactful research that can inform tailored sustainable interventions.

## INTRODUCTION

Youth living with HIV (YLHIV) face a double burden when it comes to mental health. Not only do they grapple with the challenges of navigating adolescence, but they also contend with the unique psychosocial stressors associated with HIV (Mellins & Malee, 2013; Vreeman et al., 2017). This vulnerability is particularly stark in India, where many of the children are born with HIV and face severe adversities such as loss of family members, abandonment, and economic disadvantage. Studies in India reveal a high prevalence of mental disorders among young people in the general population with approximately 7% of adolescents (13-17 years) and young adults (18-29 years) experiencing mental health challenges such as depression and anxiety. (Chadda, 2018; Murthy, 2017; Gautham et al., 2020). Among people living with HIV, rates of depression, anxiety, and psychosis can be up to 8 times higher compared to the general population, or to individuals similar in age and background but without HIV (Nedelcovych et al., 2017). Stigma, social isolation, guilt, and shame are some of the emotional burdens YLHIV carry, compounded by traumatic emotional experiences and the potential for ill health (Laryea & Gien, 1993).

Despite the vast need, comprehensively assessing the mental health of YLHIV in low-and middle-income countries (LMICs) like India, remains a significant challenge. Resource limitations are a stark reality, such as a deficit of mental health professionals comfortable with addressing concerns of YLHIV, lack of culturally adapted diagnostic tools (Gururaj et al., 2016), and the absence of integrated mental health interventions within public sector HIV service delivery models. These deficiencies hamper accurate assessment and diagnosis, leaving many YLHIV struggling with unidentified and untreated mental health problems. Discussing mental health can be challenging because of the stigma attached to it, which can lead to reluctance in seeking help or sharing experiences (Henderson et al., 2013; Pescosolido et al., 2013). A study from Karnataka State, India among a diverse socio-demographic range of the population found that the prevalence of stigma toward people with mental illness was 74.61% (Venkatesh et al., 2015). The prevailing cultural attitudes surrounding mental illness in India also perpetuate behaviors of social distancing and stigmatization among Indian youth towards individuals with mental health problems (Bell et al., 2010; Poreddi et al.,2017). The impact of such stigma is felt more intensely by younger people, who are also less likely to seek help for their mental health (Gaiha et al., 2020; Radez et al., 2021).

To begin addressing these intersectional challenges, it is often effective to intersperse those who are ‘experts by experience’ and those who conduct research. Community-Based Participatory Research (CBPR) is a powerful approach that has the potential to prioritize the voices and experiences of YLHIV (Baum et al., 2006; Cornish et al., 2023; Israel et al. 1998). It can forge equitable partnerships between those with lived experience and professional researchers and build capacity among the community (Baum et al., 2006; Cornish et al., 2023; Israel et al. 1998). This application of CBPR principles to YLHIV mental health research extends established frameworks for adult participation, while adapting them to empower YLHIV as crucial agents of change (LoIacono Merves et al., 2015).

This paper aims to describe the process and lessons learned from using a CBPR approach to develop and implement a culturally responsive mental health assessment approach among YLHIV. The two key objectives of this paper are: first, to identify actionable and contextually relevant considerations for incorporating YLHIV voices in research through CBPR principles, and second, to showcase the value of YLHIV co-creating and implementing research studies relevant to youth mental health issues.

We intentionally engaged YLHIV as ‘peer researchers’ in the research process from the very beginning, aiming to refine our tools, improve our findings, foster interventions, and enhance well-being outcomes. Peers can reduce stigma-related stress and ‘normalize’ help-seeking by openly discussing their own experiences and offering empathetic support (Sun et al., 2022). Youth involvement can also provide valuable social support and foster a positive sense of identity and group belonging (Sun et al., 2022). In addition to helping destigmatize mental health, this approach also ensures that the data collected is more authentic and reflective of the lived experiences of those facing mental health challenges.

Our approach is grounded in a human rights, social justice and equity paradigms, positioning YLHIV as research partners who have directly faced the challenges our intervention aims to address. Traditionally, these individuals are largely excluded from the research process. Children and adolescents living with HIV are typically treated as research ‘subjects’ and are often examined by ‘experts’ who lack firsthand experience about their challenges. United by the common objective of achieving social justice for this community, we actively involved these young individuals living with HIV in designing and executing a research project to pursue our goal of acquiring knowledge on the psychological and social difficulties faced by YLHIV. This approach will lend itself to designing credible and implementable solutions for effective and culturally appropriate mental health care for YLHIV.

## METHODS

### Setting

#### I’mPossible Fellowship Intervention

The mental health assessment, described in this paper, contributes to a peer-support intervention known as the I’mPossible Fellowship that was designed to empower youth living with or affected by HIV (YLHIV) in the southern states of Karnataka and Tamil Nadu. These states are among the top six states in India with the highest HIV burden. This program combines professional development, peer mentoring, and personal and academic goal pursuit to facilitate the holistic development of YLHIV. This intervention also aims to equip YLHIV with the tools to navigate resources, manage stigma, and cultivate leadership skills in vital areas like health, education, and livelihoods.

The I’mPossible Fellowship intervention is administered in collaboration with government ministries with established HIV/AIDS care initiatives and faith-based institutions in these states that provide crucial support for YLHIV by offering safe homes, medical care, educational access, and emotional support. The intervention entails connecting children and adolescent YLHIV (called ‘peers’) with experienced YLHIV mentors or ‘fellows’ for monthly one-on-one sessions. These confidential meetings provide individual support, allowing them to discuss challenges, identify gaps in resources, and develop personalized strategies for navigating health, education, and personal goals.

This mental health assessment was planned to target participants within the I’mPossible Fellowship to determine the prevalence of depression, anxiety, and feelings of stigma among recipients of the intervention (peers). The assessment findings will be subsequently used to refine the program’s mentoring and support structures, ultimately aiming to optimize mental health and resilience outcomes among YLHIV. This paper explores the YLHIV-centered engagement strategies employed during the evaluation phase of the I’mPossible Fellowship. Within this process, YLHIV peer researchers participated as ‘youth investigators’ in co-designing a mental health assessment and administering the assessment to peers.

### Selection of youth investigators

The research team identified potential youth investigator candidates from within those enrolled in or graduated from the I’mPossible Fellowship program. Candidate selection followed a purposive approach, with the following criteria: age between 18 and 24 years, living with HIV, proficiency in Kannada and English, capability to travel and attend training, and motivation for research engagement. Leveraging shared experiences and age proximity, the 18–24-year age group was chosen for their dual impact: fostering empathetic peer support and mentorship while building bridges between researchers and participants. This was anticipated to create a more comfortable research environment for participants who were either younger or the same age as the youth investigators. Proficiency in both Kannada and English was required to facilitate effective communication within the research team (mainly English speaking) and with research participants (mainly Kannada speakers). Data analysis, reporting, and dissemination required English proficiency for accurate representation of the findings, which excluded Kannada-speaking individuals with limited English. To ensure a nuanced understanding of HIV’s mental health impact, the assessment included diverse perspectives. This prompted choosing youth investigators with varied personal experiences, which included individuals living with HIV themselves, as well as those impacted through family members or caregivers living with HIV. Research staff contacted potential candidates and invited them to join the project. Six individuals meeting these criteria were chosen, consented, and officially designated as ‘youth investigators’ on the team.

### Tool Development

Early discussions led by the research team with input from two youth investigators centered around finding appropriate tools to assess well-being and mental health, as well as gauge the perspectives surrounding HIV-related stigma for YLHIV, and youth affected by HIV. After due consideration, the Patient Health Questionnaire (PHQ-9), Generalized Anxiety Disorder 7-item (GAD-7), and a modified, 4-item version of the HIV Stigma Scale (HSS) were selected for use in the mental health questionnaire. (Berger et al., 2001; Kroenke et al., 2001; Marbaniang et al., 2022; Spitzer et al., 2006).

Standardized mental health screening tools, such as the PHQ-9 and GAD-7, are often rooted in Western-defined constructs of distress, which may not be universally applicable or easily understood in non-Western cultural contexts (Kohrt et al., 2016). Acknowledging the importance of recognizing that individuals’ experience and expressions of distress are varied and culturally specific, our team considered terminologies that align with local cultural norms. This ensured that assessments are culturally relevant and minimized potential stigma associated with medicalized language and standardized assessments (Desai & Chaturvedi, 2017; Kohrt et al., 2016).

Several rounds of discussion were conducted to deeply review these screening instruments. Input was obtained from youth investigators who addressed comprehension challenges and wording issues in standardized tools (PHQ-9, GAD-7, and HIV Stigma Scale). They identified and suggested improvements for clarity and usability using role play and informal focus group discussions.

One example is in the PHQ-9, which uses polychotomous statements like “Trouble falling or staying asleep or sleeping too much.” The youth investigators identified potential comprehension issues with such statements; participants might compartmentalize the options, potentially overlooking the possibility of experiencing both or focusing solely on one symptom. Through discussions of alternative approaches, the youth investigators proposed splitting the statement into two separate questions: “Do *you have trouble falling asleep?”* and “*Do you sleep too much?”*. This clearer format reduced the risk of participants accidentally overlooking one aspect.

Furthermore, despite pre-existing Kannada translations for all survey tools, the youth investigators recognized the linguistic diversity within Kannada dialects across districts, identifying specific words that were not contextually appropriate and suggesting alternatives to enhance comprehension. One such word was “*fidgety*”, used in the PHQ-9 that had no direct Kannada translation and may be interpreted in multiple ways. Therefore, it was crucial to identify the appropriate Kannada term that encompassed the intended meaning for the assessment. After multiple rounds of review, the final versions were back-translated into English and reviewed by the senior investigators to ensure fidelity to the purpose of the mental health screening aim.

### Training

Selected youth investigators completed the Johns Hopkins Bloomberg School of Public Health Basic Human Subjects Research Protection CITI program course and a two-day, eight-session mental health questionnaire training program. This training program was designed to provide them with background knowledge of mental health and comprehensive preparation for administering the mental health questionnaire comprising the PHQ-9, GAD-7, and abbreviated HIV Stigma Scale. Training sessions encompassed tool significance, interview techniques, communication skills, and addressing challenges. They featured live demonstrations, group practice, and practical evaluations to ensure effective scale administration with attention to well-being, cultural sensitivity, and ethics (see Appendix 1). Following the initial pedagogic sessions, training techniques included interactive discussions, role play, and debate. At the end of the sessions, the youth investigators filled out a feedback form on their readiness to administer the mental health questionnaire and shared their overall experience.

### Peer-Led Mental Health Assessment Process

Following assessment of their readiness by the research team, the youth investigators conducted field mental health assessments in various urban and rural settings across Karnataka and Tamil Nadu. The study enrolled 206 participants (8-25 years old), including YLHIV and youth affected by HIV due to a family member or caregiver’s diagnosis. Youth investigators established rapport with the participants in the assessment (i.e., peers who received the I’mPossible fellowship intervention), explained the questionnaire’s purpose, assured confidentiality, inquired about questions or concerns, and obtained consent/assent to proceed with the questionnaire. They completed assessments based on participants’ responses, and ensured ethical standards, participant well-being, and sensitivity throughout the process. A professional counselor was available during all these sessions. If a participant had responses that indicated significant immediate risk, the youth investigator promptly connected them with the counselor, either in-person or by phone. Following these field assessments, youth investigators had regular debriefing sessions with the research team to address any challenges or concerns that emerged during the questionnaire administration process.

### Reflective essays

Following the administration of a minimum of ten mental health assessments to their peers, the youth investigators were tasked with composing reflective essays. These essays centered on their own experiences in administering the mental health questionnaires in the field and their preparedness for data collection, offering insights and reflections on the entire process, from developing the tools, training, and administration. They were asked to consider the following prompts in their reflection (1) Describe their overall experience participating in the training program. (2) Identify key learnings or insights they gained from the training. (3) Discuss any challenges they encountered while administering the mental health questionnaires. (4) Reflect on their level of preparedness to conduct assessments confidently and effectively. Each essay was closely reviewed by the research team. Recurring themes, patterns, significant quotes, and emotion expressed were noted and coded accordingly. Potential themes were constructed to identify commonalities, and then refined to ensure they were distinct and internally coherent. The findings were then triangulated with other data sources, such as the training feedback form and researcher observations, providing corroborating evidence for the emergent themes.

## LESSONS LEARNED

The CBPR process we applied to conducting a mental health assessment among YLHIV offers many lessons which are summarized in this report. Overall, the CBPR approach enhanced not only the quality of research but also nurtured competencies and personal growth. Monitoring modes such as training session feedback forms, reflective essays, and regular debriefing sessions allowed for the identification of benefits, challenges, and areas requiring additional support. The key takeaways from this approach included the incorporation of interactive training, equitable research, secure environment for peers, and youth leadership capacity building.

### Training that is interactive and enriching

Interactive demonstrations and practice sessions with clear explanations significantly improved comprehension and confidence. Initial training sessions for the screening tools (PHQ-9, GAD-7, HSS) relied heavily on a lecture format. A majority of youth investigators provided feedback that they preferred a more engaging approach. Utilizing interactive approaches during the latter parts of the training helped to ensure understanding and skill development. Youth investigators felt that the training effectively utilized interactive demonstrations and peer collaboration to facilitate understanding and engagement. Specific elements like role play, interview practice with peers, and explanation through demonstrations strongly resonated with participants. The reflective essays revealed that the practice sessions introduced effective strategies for handling diverse situations that might arise during the administration process. As a result, the youth investigators felt more at ease in creating a safe space for the participants. One youth investigator wrote in their essay,

“*We are benefiting from the practice [sessions]. This way really helps create a supportive environment and customize sessions for each child. This class was very valuable to me, and I learned how to handle different people*.*” [Youth Investigator 1]*

### A research environment that is collaborative and equitable

Encouraging peer discussions, joint research efforts, and regular feedback loops helped to build a collaborative research environment and leverage collective knowledge. In the training feedback form, all youth investigators indicated they felt empowered to engage in research discussions and joint efforts with senior researchers. The act of sharing and receiving support also fostered a sense of community, allowing youth investigators to rely on each other and the research team, and experiencing respectful guidance and encouragement. Furthermore, hearing how others handled various situations instilled confidence in their own abilities and provided practical models for successful survey administration. Thus, creating a culture of collaborative problem-solving, shared decision-making and mutual co-learning fostered a supportive community and enhanced research quality. One youth investigator described their experience, stating,

*“On the first day of training, it was difficult to understand the tools, but on the second day, with the help of a demo done by [the research coordinators], it made it easy to understand. Along with the demo, we in the group tried asking one another questions, and this made us comfortable asking the students in a better way*.*” [Youth Investigator 4]*

### Creation of safe spaces for vulnerability

Another significant outcome of youth involvement in administering mental health assessments was their capacity to establish a secure and inclusive environment for participants to openly express their emotional experiences. This outcome reflects the far-reaching impact of youth engagement in research, demonstrating that beyond the collection of data, their involvement led to open and candid conversations with their peers, ultimately promoting enriched social connectedness among young individuals grappling with similar circumstances and challenges. These open conversations can potentially trigger emotional responses in both the youth investigators and the participants. Participants might experience distress or discomfort when addressing sensitive topics pertaining to their mental health, which may be alleviated with the presence of researchers with similar lived experiences.

One youth investigator, when reflecting on their experience in administering the mental health questionnaire to peers, stated:

*“These students were really open and shared their feelings and worries with the leaders of their group. They felt comfortable talking to us and told us about their lives and the things that are hard for them*.*” [Youth Investigator 4]*

The presence of on-site counselors played a crucial role in ensuring the well-being of both participants and youth investigators. Participants had the opportunity to engage in post-assessment debriefing with a professional and could be promptly referred for crisis intervention when needed. Youth investigators also had the option to seek guidance from the counselor when they encountered uncertainties or challenges during the assessment. Integrating mental health training for YLHIV researchers alongside on-site counselors equipped them with skills to handle emotional responses and provide appropriate support.

### Youth involvement that builds confidence and leadership

By prioritizing capacity building through targeted support and fostering empathy, youth investigators felt empowered to contribute to valuable research and develop leadership skills. Many highlighted their enhanced ability to connect with their peers on a deeper level. Some expressed uncertainty about handling emotional and sensitive topics. In response, the trainers implemented additional confidence-building exercises to provide targeted support for navigating sensitive subjects and offered opportunities for skill development.

In their reflective essays, youth investigators shared personal transformations, highlighting newfound confidence and communication skills. One youth investigator stated:

*“Before I joined this training, I did not have self-confidence and [had] stage fright. When I attended this training, I learned to be confident in doing some activities, surveys… [Youth Investigator 2]”*

Another youth investigator, reflected on the training process, emphasizing its multifaceted significance, by stating:

*“This extensive training not only equipped us with the necessary skills to guide the children through the survey, but also underscored the significance of building our own confidence as instructors. [Youth Investigator 3]”*

## DISCUSSION

Given the scarcity of widely publicized models for health-related youth CBPR in India, especially with vulnerable populations like YLHIV, this project stands as a crucial case study and offers valuable lessons learned. This project operationalized CBPR principles within YLHIV mental health assessment, offering not only valuable insights into YLHIV mental health but also a potential template for other research studies on complex issues regarding youth (Table 1).

**Table 1.** Community-Based Participatory Research Principles for YLHIV Mental Health Assessment.

| Principle | Description |
| --- | --- |
| 1. Recognizing YLHIV as a distinct community | Mental health assessments are tailored to YLHIV needs, valuing their shared experiences and cultural contexts. |
| 2. Building on strengths and resources | YLHIV are empowered to contribute their expertise and perspectives utilizing their inherent resilience and existing knowledge of the community. |
| 3. Equitable partnerships | Power imbalances are broken down as YLHIV actively participate in research design, data collection, and data interpretation, leading to shared decision-making and mutual co-learning. In doing so, YLHIV take ownership of their own mental health narratives. |
| 4. Actionable knowledge | Research findings translate into concrete interventions and culturally appropriate mental health programs that may be co-created by YLHIV. |
| 5. Co-learning and empowerment to address social inequities | Researchers and YLHIV engage in a reciprocal exchange of knowledge and expertise. YLHIV gain skills and confidence to advocate for their own needs and build capacity within the community. |
| 6. Cyclical and iterative process | Ongoing feedback from YLHIV ensures assessments remain relevant and responsive to their evolving needs, fostering a dynamic and adaptable research process. |
| 7. Holistic perspective | Moving beyond symptoms to acknowledge the social, economic, and environmental factors impacting YLHIV's mental health, leading to interventions that address the root causes of their vulnerabilities. |
| 8. Transparent and respectful knowledge sharing | Research findings will be openly shared by YLHIV, researchers, and community partners, empowering all stakeholders to utilize the knowledge. |

Adapted from Israel et al, 1998.

Recognizing YLHIV as a distinct community ensures that researchers acknowledge and value their unique experiences and cultural contexts (Israel et al., 1998). YLHIV, in this study, incorporated cultural and identity considerations during tool development that ensured that the screening instruments resonated with their lived experiences. With the goal of achieving contextual relevance, Mazzuca et al. (2018) modified the PHQ-9 by revising conflicting statements, incorporating local idioms, and adding culturally specific examples to enhance the tool’s utility for low caste adolescent girls in rural south India. While it is crucial to adapt these Western-construct-based tools to reflect the local meanings and experiences of the communities being assessed, simply relying on translation and back-translation of these instruments’ risks misinterpretation and errors (Mazzuca et al., 2018; Parkerson et al.,2015).

In our CBPR approach, we not only modified mental health screening tools based on feedback from YLHIV, we also built upon their inherent strengths and resources by having them administer the mental health questionnaire. Youth investigators created a comfortable research environment for peers to answer the questions honestly through open communication and active listening. Understanding the unique challenges of youth and building upon their range of competencies while engaging them as research partners provides a valuable approach to youth health intervention research (LoIacono Merves et al., 2015). It is known that YLHIV often face significant employment barriers, including health-related concerns, limited job options, and persistent fear of discrimination (Maulsby et al., 2020). Participating in this partnership equipped youth investigators with relevant vocational skills for the first time, such as report writing, public speaking, and engaging with a diverse group of people. Such skills are easily transferable and valuable to future employment.

Despite CBPR being fundamentally built on equitable partnerships, a concerning gap exists in youth CBPR research: the underrepresentation of genuine youth partnerships (Jacquez et al.,2013). Even in studies with youth involvement, the extent of their participation varies considerably (Jacquez et al.,2013). In this study, YLHIV youth investigators worked with research staff and demonstrated a desire to be actively engaged in every aspect of the research process (from tool selection to tool administration, and ongoing data management, analysis, and dissemination). The success of this partnership was dependent on making intentional individual connections between the youth and adult investigators, that utilized motivational language and personalized support (LoIacono Merves et al., 2015). Due to the potential for emotionally charged conversations that could be triggering, it was crucial for research staff and counselors to provide a judgment-free space for the youth investigators to process their thoughts and feelings. Although checking in with young investigators is not explicitly an expectation of CBPR, it is still beneficial to keep youth interested and committed long-term (LoIacono Merves et al., 2016). Engaging youth appropriately and respectfully was key to leveraging their expertise and ensuring equitable research relationships.

This study is not without limitations. First, our approach drew heavily on the perspectives of youth investigators and did not include data directly from YLHIV participants in addition, which may have provided a more comprehensive understanding of the assessment experience. We plan to include seeking the participant component in our next iteration of the CBPR approach. Second, by including only those youth investigators with dual proficiency in English and Kannada, there was a limitation in inclusivity of others in the community with limited knowledge of English but with rich experience and motivation to be part of the research community. We plan to address this by utilizing better translation services and expanding research capacity within the community. Finally, while this study focused on establishing robust data generation for YLHIV mental health research, it acknowledges limitations in applying CBPR principles during the analysis and dissemination stages both of which are currently in the planning phases. Therefore, though this study recognizes the critical role of CBPR principles, the integration of the principles of holistic perspectives and transparent knowledge sharing is yet to be implemented. In this ongoing activity, we intend to intentionally engage youth investigators in these stages by (i) actively soliciting their interpretation of analytic findings, and by (ii) creating opportunities for them to take an active role in manuscript preparation, be primary presenters at conferences, and create lay materials for dissemination to the larger YLHIV community. Future activities with youth investigators will include co-creating interventions informed by the mental health assessments to better support the psychosocial well-being of YLHIV participants.

## CONCLUSION

Using CBPR principles, youth within the local HIV community in Karnataka and Tamil Nadu harnessed evidence-based tools to deliver culturally tailored mental health screening instruments. They have not only played an instrumental role in developing these tools but have also efficiently administered them to assess the mental health status of their own community of YLHIV in the respective regions. This CBPR process will proceed to include the youth investigators in data analysis and dissemination of findings. The next phase will involve collaborating with young individuals to co-create peer-led interventions that can enhance the delivery of mental health services within the community. This study serves as an illustrative model of involving youth in mental health research and highlights the valuable insights and lessons learned, encouraging fellow researchers to incorporate the perspectives and experiences of young individuals for overall enhanced research outcomes.

## Supporting information

Appendix 1. Summary of Group Training Sessions

## Data Availability

All data produced in the present work are contained in the manuscript.

