## Appendix 1. Summary of Group Training Sessions for "Partnering for Progress: Lessons Learned from a Mental Health Assessment for Youth Living with HIV in India through Community-Based Participatory Research"

| Session | Description | Activities |
|---|---|---|
| **Day 1** | | |
| 1 | Introduction and Orientation (45 minutes) | • Welcome the youth researchers and provide an overview of the training agenda.<br>• Explain the purpose of the study and the significance of administering the PHQ-9, GAD-7, and HIV Stigma Scale accurately and sensitively.<br>• Discuss the ethical considerations and the importance of confidentiality and informed consent.<br>• Provide a detailed explanation of the PHQ-9, GAD-7, and HIV Stigma Scale.<br>• Go through each question in the scales, explaining their meaning and how to interpret the responses (e.g., Likert scale - strongly disagree to strongly agree). |
| 2 | Understanding the Scales (45 minutes) | • Present examples of relevant studies where these scales were used and how they contributed to the research.<br>• Showcase a study that employed the HIV Stigma Scale to measure stigma experienced by people living with HIV in a particular region.<br>• Emphasize the importance of cultural sensitivity and understanding. |
| **Day 2** | | |
| 3 | Demonstration (45 minutes) | • Facilitators will conduct a demonstration of how to administer the PHQ-9, GAD-7, and HIV Stigma Scale to a mock participant.<br>• The demonstration will showcase the correct approach, demonstrating sensitivity and respect towards the participant's feelings and experiences.<br>• Researchers will observe the demonstration and take note of the proper techniques and communication skills used during the assessment. |
| 4 | Interview Techniques and Communication Skills (30 minutes) | • Train the youth researchers in effective interview techniques, such as active listening, empathy, and non-judgmental attitudes.<br>• Teach them how to ask open-ended questions to encourage participants to share their responses more freely. |
| 5 | Handling Difficult Situations (20 minutes) | • Discuss potential challenging scenarios during interviews, such as emotional distress or disclosure of sensitive information.<br>• Provide guidance on how to offer support, express empathy, and refer participants to mental health professionals if necessary. |
| 6 | Group Practice and Discussion (45 minutes) | • Youth researchers engage in hands-on group practice to improve their interview skills in administering the PHQ-9, GAD-7, and HIV Stigma Scale.<br>• They are divided into groups of three, taking turns as the interviewer, interviewee (role-playing as a young person living with HIV), and observer.<br>• After the practice rounds, researchers discuss their experiences and receive feedback from facilitators to enhance their communication and interview techniques. |
| 7 | Data Recording and Documentation (5 minutes) | • Instruct the youth researchers on proper data recording procedures.<br>• Emphasize the importance of accurate and complete data collection to ensure reliable results. |
| 8 | Debriefing, Support, and Role of Youth Researchers and Conclusion (10 minutes) | • Schedule regular debriefing sessions for the youth researchers to discuss their experiences, share challenges, and seek support.<br>• Encourage peer support and collaboration among the researchers.<br>• Remind the youth researchers of their essential role as ambassadors of the study and the impact of their work on research outcomes and participant well-being.<br>• Provide final instructions and reminders for the data collection process.<br>• Conclude the training and express appreciation for their participation. |
